# Investigation and analysis of blood-borne occupational exposure of medical staff in a hospital in Hubei Province

**DOI:** 10.1101/2021.06.07.21258449

**Authors:** Yan Yang, Junzheng Yang

**Affiliations:** Shiyan people’s hospital (Hubei Medical College Affiliated People’s Hospital), Shiyan, Hubei, China, 442000; Guangzhou Packgene biotechnology Co., Ltd, Guangzhou, Guangdong, China, 510000

**Keywords:** blood-borne occupational exposure, medical staff, safety and protection

## Abstract

**Objectives:** By collecting and sorting out the data of 220 blood-borne occupational exposure medical staff’ in a hospital in Hubei Province, investigated the present situations and problems of the blood-borne occupational exposure, and the causes were analyzed and the improvement measures for personal safety of the medical staff were also made, which may provide guidance and reference significance for the medical staff.

**Methods:** a retrospective survey was conducted to analyze occupational exposure populations by influence factors (gender, age, occupation, occurrence places, occurrence links, and exposure of pathogens types.

**Results:** There were 220 medical staff of blood-borne occupational exposure in the hospital in Hubei Province from 2015 to 2017 to be investigated, and the blood-borne occupational exposure populations were mainly concentrated on nurses (65%) and doctors (31.82%) (both of populations accounted for 96.82% in total occupational exposure medical staff), and the age of blood-borne occupational exposure populations were mainly 20~29 years old (61.82%), followed by 30~39 years old (26.82%), 40~49 years old (8.64%), and >50 years old (2.73%). The main occurrence places for blood-borne occupational exposure were wards (46.36%) and treatment room/disposal rooms(26.36%); the main occurrence links of blood-borne occupational exposure were mainly injured during needle extraction (29.09%), stab and cut during treatment (23.64%), disposal of waste (17.27%) and accidental injury during operation (16.82%). The main pathogen types of blood-borne occupational exposure were hepatitis B (HBV) and syphilis, accounting for 36.36% and 10.45%, respectively.

**Conclusions:** The population of blood-borne occupational exposure in this hospital mainly concentrated in nurses aged 20-29 years old, followed by doctors, mainly happened at wards and treatment rooms/disposal rooms; the high-risk links were mainly injured during needle extraction and stab and cut during treatment, and the main pathogen types were HBV and syphilis. Those evidences showed that to strengthen the awareness of prevention and operation standard training for the related medical staff (mainly nurses and doctors aged 20-29 years old) and to supervise the relevant departments to do a good job of supervision, it can greatly reduce the incidence and risk of blood-borne occupational exposure in this hospital.

## 1. Introduction

Blood-borne occupational exposure of medical staff refers to a kind of occupational exposure in which medical staff are exposed to toxic, harmful substances or pathogens of infectious diseases in the process of diagnosis, treatment and nursing, thus damaging their health or endangering their lives. Infectious diseases caused by blood-borne occupational exposure not only affect the physical and mental health of medical staff, cause the loss of health resources, but also can spread for the second time, causing harm to the family of medical staff. In recent years, it has become a big problem faced by medical staff, and has been paid more and more attention by hospital administrators and researchers^[1]^. Since the introduction of Xinglin hospital infection monitoring software in 2014, the hospital in Hubei province has started to systematically monitor blood-borne occupational exposure, which could summarize the causes and population of blood-borne occupational exposure of hospital staff, and provide the basis for taking relevant measures to reduce the risk of blood-borne occupational exposure of hospital staff. In this paper, we reported and summarized the problems of blood-borne occupational exposure of medical staffs in a hospital from 2015 to 2017 according to the data collected by Xinglin hospital infection monitoring software, sorted out the data according to gender, age, occupation, occurrence places, occurrence links and pathogen types, so as to put forward corresponding measures to reduce the occurrence of occupational exposure and effectively protect the occupational safety of medical staff.

## 2. Methods

### 1.1 Investigation objects

The objects of this study were the medical staff who reported blood-borne occupational exposure to the infection management department in the hospital in Hubei Province from 2015 to 2017, including doctors, nurses, technicians, service workers (cleaners, interns and trainees).

### 1.2 Investigation methods

Using the pre-designed registration form of blood-borne occupational exposure in Xinglin software, the blood-borne occupational exposure situation reported in the hospital in Hubei Province from 2015 to 2017 was analyzed retrospectively, including the basic personal information (age, gender, occupation, working years, etc.) and the related situation of occupational exposure (exposure time, exposure place, exposure mode, post exposure treatment, tracking, etc.).

## 2. Results

### 2.1 Gender distribution, occupation distribution and age distribution of blood-borne occupational exposure population in the hospital in Hubei Province

A total of 220 cases of blood-borne occupational exposure were reported in the hospital in Hubei Province from 2015 to 2017. According to gender, 54 cases (24.55%) were male and 166 cases (75.45%) were female in blood-borne occupational exposure in the hospital (Table 1); According to occupation, 143 cases (65%) were nurses in blood-borne occupational exposure in the hospital, followed were doctors (70 cases, 31.82%) (Table 2); the data showed that the age 20-29 years people were 136 cases (61.82%) in blood-borne occupational exposure, followed by aged 30-39 years (59 cases, 26.82%), aged 40-49 years (19 cases, 8.64%) and aged⩾ 50 years (19 cases, 2.73%) (Table 3).

**Table 1.** Gender distribution of occupational exposure population in medical staff in the hospital

| gender | cases | proportion (%) |
| --- | --- | --- |
| Male | 54 | 24.55 |
| Female | 166 | 75.45 |
| Total | 220 | 100 |

**Table 2.** Occupation distribution of occupational exposure population in medical staff in the hospital

| occupation | cases | proportion (%) |
| --- | --- | --- |
| Nurse | 143 | 65 |
| Doctor | 70 | 31.82 |
| Technician | 4 | 1.82 |
| Service worker | 3 | 1.36 |
| Total | 220 | 100 |

**Table 3.**
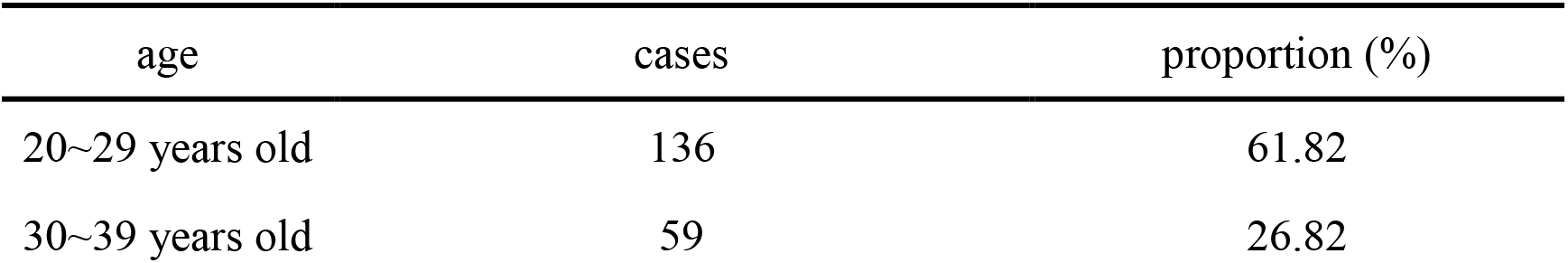
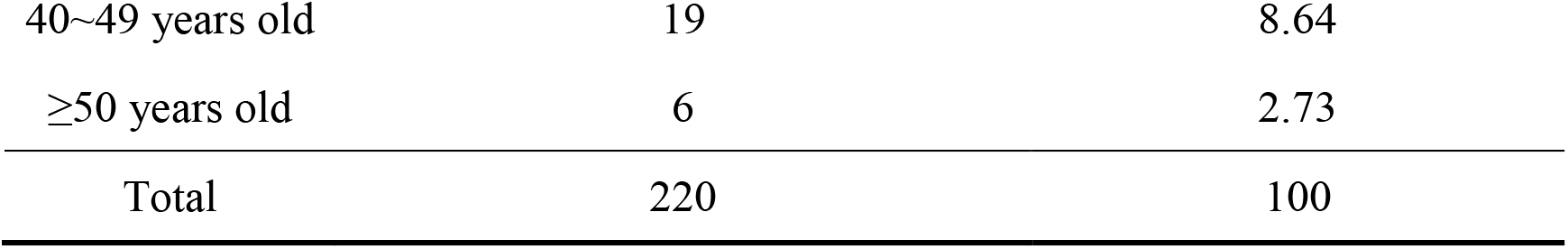
Age distribution of occupational exposure population in medical staff in the hospital

### 2.2 Occurrence place and occurrence link distribution of blood-borne occupational exposure in medical staff in the hospital

The data showed that the main occurrence places of occurrence in blood-borne occupational exposure were wards (102 cases, 46.36%), treatment room/disposal room (58 cases, 26.36%) and operation room (46 cases, 20.91%), this three kinds of occurrence places accounted for 93.64% of the total number of blood-borne occupational exposure populations in medical staff in the hospital (Table 4); It showed that the occurrence links of blood-borne occupational exposure were mainly concentrated in injured during needle extraction (64 cases, 29.09%), stab and cut during treatment (52 cases, 23.64%), disposal of waste (38 cases, 17.27%), accidental injury during operation (37 cases, 16.82%), splash or body surface contact (20 cases, 9.09%)(Table 5).

**Table 4.**
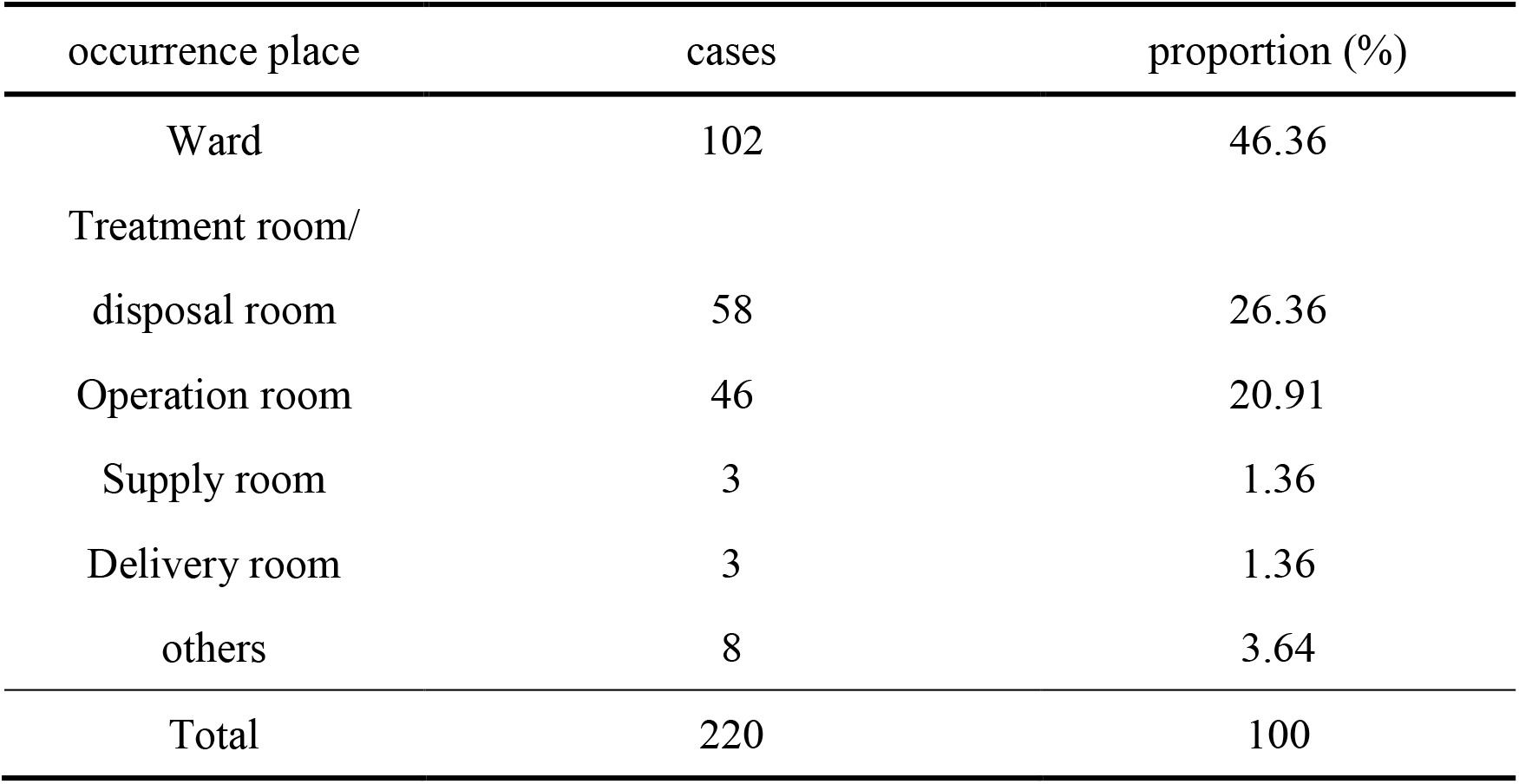
Occurrence places distribution of blood-borne occupational exposure population in medical staff in this hospital

| occurrence place | cases | proportion (%) |
| --- | --- | --- |
| Ward | 102 | 46.36 |
| Treatment room/<br>disposal room | 58 | 26.36 |
| Operation room | 46 | 20.91 |
| Supply room | 3 | 1.36 |
| Delivery room | 3 | 1.36 |
| others | 8 | 3.64 |
| Total | 220 | 100 |

**Table 5.** Occurrence link distribution of blood-borne occupational exposure population in medical staff in this hospital

| occurrence link | cases | proportion (%) |
| --- | --- | --- |
| Injured during needle extraction | 64 | 29.09 |
| Stab and cut during treatment | 52 | 23.64 |
| Disposal of waste | 38 | 17.27 |
| Accidental injury during operation | 37 | 16.82 |
| Splash or body surface contact | 20 | 9.09 |
| When cleaning instruments | 9 | 4.09 |
| Total | 220 | 100 |

### 2.3 Pathogen types distribution of blood-borne occupational exposure population in medical staff in this hospital

In pathogen types distribution statistics, there were a lot of medical staff after blood-borne occupational exposure was not monitored/unclear, accounting for 39.55%, followed by HBV and syphilis, accounting for 36.36% and 10.45%, respectively. The details were shown in Table 6. The statistics results demonstrated that the hospital administrators did not have enough understanding of blood-borne occupational exposure of medical staff, and did not take necessary pathogen examination for medical staff after blood-borne occupational exposure, reduce the harm of blood borne occupational exposure to medical staff.

**Table 6.**
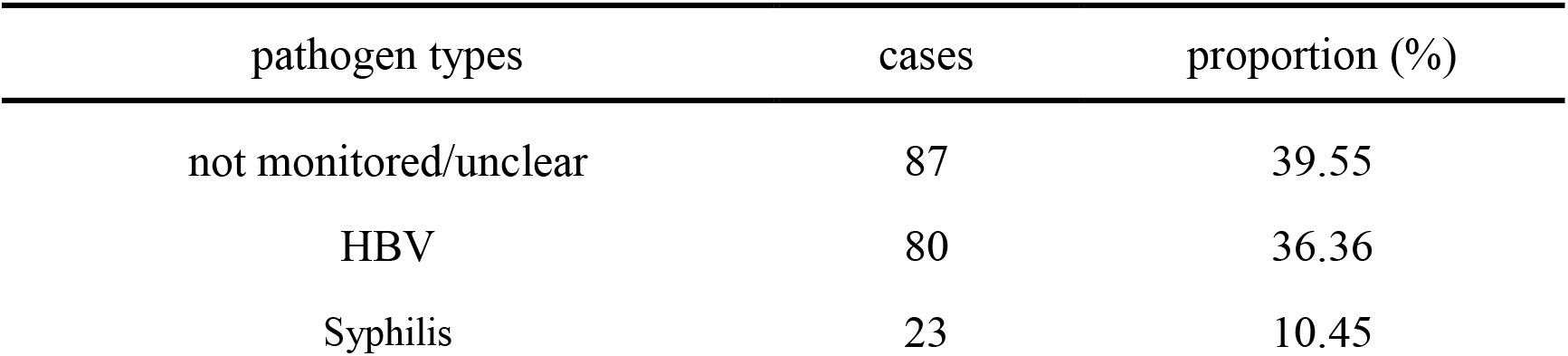
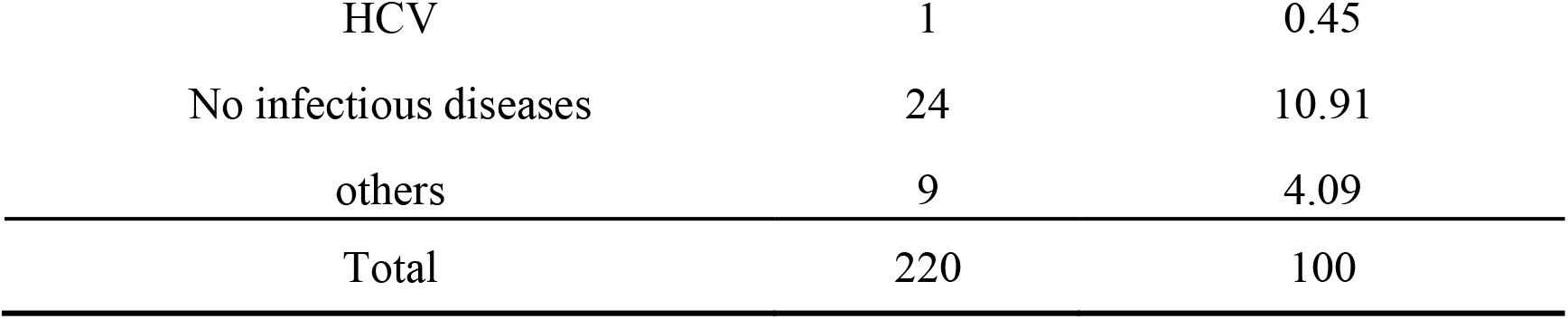
Pathogen types distribution of blood-borne occupational exposure population in medical staff in this hospital

### 2.4 Therapeutic method after blood-borne occupational exposure

220 cases of blood-borne occupational exposure medical staff were treated by washing and disinfection treatment, the disinfection treatment rate was 100%, and 149 cases of occupational exposure were treated and checked in the hospital, and the preventive drug use rate was 67.73%. No infection was found after treatment.

## 3 Discussion

Medical staff in the hospital are faced with greater occupational exposure risks due to their special work environment, and blood-borne occupational exposure is the most common, and poses a great threat to the normal life and physical and mental health of medical staff^[2]^. It is necessary to strengthen the investigation and analysis of occupational exposure population, and strive to start from the exposure sources and high-risk links, To minimize the incidence of blood-borne occupational exposure of medical staff^[3]^. In this hospital in Hubei Province, the real-time report was made through the introduction of hospital monitoring system; carry out regular training on occupational protection and safety in the hospital; The hospital is responsible for the diagnosis and treatment of occupational safety protection incidents in the normal working hours, so as to provide security for medical staff.

The statistics results showed that nurses were main populations in the blood-borne occupational exposure population, accounting for 65%, which was consistent with the results of relevant domestic reports^[4–5]^. This is because nurses had to carry out a lot of injection, infusion, blood collection and other operations every day, they have more opportunities to contact syringes, infusion sets and other sharp instruments, the probability of blood-borne occupational exposure is also higher than that of other occupations. The young medical staff aged 20-29 years old is the main population with the most occupational exposure, they had short working years, inexperienced, unskilled operation, and sometimes not according to the standard operation, which is more prone to accidental sharp injuries at work. On the one hand, the occurrence of blood-borne occupational exposure is related to the medical staff’s poor awareness of prevention, poor clinical operation standard and low compliance with standard prevention^[6]^; On the other hand, it is closely related to poor management and failure to implement effective supervision. Therefore, it is suggested that we should continue to strengthen the training of occupational safety for medical staff, especially for new employees, and strengthen the management at hospital and department levels. Full time personnel should go deep into clinical supervision regularly to improve the personal protection awareness of medical staff.

The results showed that the most common link of occupational exposure was when nurses took blood and pulled out the needle after infusion, timely and safely put the pulled-out needle into the sharp tool box, which could effectively prevent the occurrence of stab injury; It is also an important reason for occupational contact to return the needle cap with both of hands, the chance of returning the needle cap should be reduced, and the single glove needle cap should be used when necessary^[7]^. The accidental injuries in the operating room are also the places and links with more occupational exposure in the hospital, which mainly occur in the process of operation. It reminds doctors to be calm and take personal protective measures when facing patients, especially infectious patients; the curved plate must be used to transfer sharp instruments such as knife and scissors during the operation, not by naked hand; The materials should be placed in appropriate containers in time to avoid injury to others after use; every personnel should cooperate with each other to avoid mistakes in busy work.

In addition, the pathogen detection of the blood-borne occupational exposure showed that nearly 40% of the patients were not detected before treatment or operation. For this reason, medical staff were required to conduct a full set of blood transfusion examination before treatment on the basis of standard prevention, so as to determine whether they were patients with infectious diseases, so as to take targeted prevention and control measures. Among the known pathogens, HBV and syphilis are the most common pathogens, which is also related to the number of people infected with such pathogens in the social population. It suggests that we should focus on strengthening the standard prevention of such infectious diseases and post-treatment of occupational exposure in training, and it is suggested that those who have the conditions should take the initiative to vaccinate against HBV. After the occurrence of exposure, do a good job with local treatment, timely and actively report to the infection management department, and give preventive measures according to the exposure situation, which can effectively control the occurrence of infection after occupational exposure and protect the occupational safety of medical staff.

Through the analysis of blood-borne occupational exposure of medical staff in the hospital in Hubei Province, the main occupational exposure population, exposure place and high-risk occurrence link of occupational exposure were found out, which provided the basis for the next work of occupational safety protection. Medical institutions should establish and improve the relevant system of occupational protection, strengthen training, improve the self-protection awareness and treatment skills of medical staff, and provide a good occupational environment for medical staff by regular physical examination and vaccination.

## Data Availability

The data used to support the findings of this study are available from the corresponding author upon request.

## 4. Acknowledgement

None.

